# Development of the PSYCHS: Positive SYmptoms and Diagnostic Criteria for the CAARMS Harmonized with the SIPS

**DOI:** 10.1101/2023.04.29.23289226

**Authors:** Scott W. Woods, Sophie Parker, Melissa J. Kerr, Barbara C. Walsh, S. Andrea Wijtenburg, Nicholas Prunier, Angela R. Nunez, Kate Buccilli, Catalina Mourgues-Codern, Kali Brummitt, Kyle S. Kinney, Carli Trankler, Julia Szacilo, Beau-Luke Colton, Munaza Ali, Anastasia Haidar, Tashrif Billah, Kevin Huynh, Uzair Ahmed, Laura L. Adery, Cheryl M. Corcoran, Diana O. Perkins, Jason Schiffman, Jesus Perez, Daniel Mamah, Lauren M. Ellman, Albert R. Powers, Michael J. Coleman, Alan Anticevic, Paolo Fusar-Poli, John M. Kane, Rene S. Kahn, Patrick D. McGorry, Carrie E. Bearden, Martha E. Shenton, Barnaby Nelson, Monica E. Calkins, Larry Hendricks, Sylvain Bouix, Jean Addington, Thomas H. McGlashan, Alison R. Yung, the Accelerating Medicines Partnership Schizophrenia, AMP SCZ Working Group and Subgroup leaders (not previously listed):, Kelly Allott, Scott R. Clark, Tina Kapur, S. Lavoie, Kathryn E. Lewandowski, Daniel H. Mathalon, Ofer Pasternak, William S. Stone, John Torous, National Institute of Mental Health Project Scientists:, Laura M. Rowland, Ming Zhan, Research Network and DPACC Investigators (not previously listed):, Paul Amminger, Celso Arango, Matthew R. Broome, Kristin S. Cadenhead, Eric Y.H. Chen, Jimmy Choi, Kang Ik Kevin Cho, Philippe Conus, Barbara A. Cornblatt, Louise Birkedal Glenthøj, Leslie E. Horton, Joseph Kambeitz, Matcheri S. Keshavan, Nikolaos Koutsouleris, Kerstin Langbein, Covadonga Martinez Diaz-Caneja, Vijay A. Mittal, Merete Nordentoft, Pablo A. Gaspar Ramos, Godfrey D. Pearlson, Jai L. Shah, Stefan Smesny, Gregory P. Strauss, Jijun Wang, Study Coordinators and Project Managers (not previously listed):, Patricia J. Marcy, Priya Matneja, Christina Phassouliotis, Susan Ray, Collum Snowball, Jessica Spark, Sophie Tod, Individual names of AMP SCZ collaborators are listed in the Acknowledgment

## Abstract

**Aim:** To harmonize two ascertainment and severity rating instruments commonly used for the clinical high risk syndrome for psychosis (CHR-P): the Structured Interview for Psychosis-risk Syndromes (SIPS) and the Comprehensive Assessment of At-Risk Mental States (CAARMS).

**Methods:** The initial workshop is described in the companion report from Addington et al. After the workshop, lead experts for each instrument continued harmonizing attenuated positive symptoms and criteria for psychosis and CHR-P through an intensive series of joint videoconferences.

**Results:** Full harmonization was achieved for attenuated positive symptom ratings and psychosis criteria, and partial harmonization for CHR-P criteria. The semi-structured interview, named <u>P</u>ositive <u>SY</u>mptoms and Diagnostic Criteria for the <u>C</u>AARMS <u>H</u>armonized with the <u>S</u>IPS (PSYCHS), generates CHR-P criteria and severity scores for both CAARMS and SIPS.

**Conclusion:** Using the PSYCHS for CHR-P ascertainment, conversion determination, and attenuated positive symptom severity rating will help in comparing findings across studies and in meta-analyses.

## 1 INTRODUCTION

The clinical high-risk syndrome for psychosis (CHR-P), also known as the ultra-high risk (UHR) mental state, was first described 25 years ago (Yung et al, 1996) and has provided an influential paradigm for early detection and intervention in psychosis. CHR-P syndrome patients are youth and young adults who are symptomatic and impaired and also at risk for developing frankly psychotic disorders (Woods et al, 2021; Woods et al, 2001). The condition is listed in DSM-5 as Attenuated Psychosis Syndrome (American Psychiatric Association, 2022) as one of four specified “Other Specified Schizophrenia Spectrum and Other Psychotic Disorders” (ICD-10 F28) under the construct of “Conditions for Further Study”; further study has suggested substantial validity (Mensi et al, 2021; Salazar de Pablo et al, 2020). CHR-P syndromes are associated with a meta-analytic 20% probability of developing psychosis at two years, which increases over the long term peaking to 35% at 10-years (de Pablo et al, 2021b). Most CHR-P individuals who will not develop psychosis will continue displaying other poor mental health outcomes at follow-up (Addington et al, 2019; de Pablo et al, 2021a). Multiple biological markers predict onset of psychosis in CHR-P patients (Fusar-Poli et al, 2020), including recent evidence that thinning of cerebral cortex precedes and predicts psychosis (Collins et al, 2022). CHR-P a common, if under-recognized, condition, as evidenced by meta-analytic estimates of point prevalence in the general youth population (1.7%) and in the population of youth presenting for psychiatric care (19.2%) (Salazar de Pablo et al, 2021). A recent bibliographic analysis identified 1,637 unique research data publications, with two or more publications originating from 1,573 separate institutions in 49 countries (Lee et al, 2022). More than 100 specialty clinics for CHR-P have been organized in multiple countries across six continents (Kotlicka-Antczak et al, 2020).

Two semi-structured interviews have commonly been used to ascertain patients for CHR-P and to rate their severity of illness over time (Andreou et al, 2019; Daneault et al, 2013; Olsen and Rosenbaum, 2006): the Structured Interview for Psychosis-risk Syndromes (SIPS) and the Comprehensive Assessment of At-Risk Mental States (CAARMS) (Miller et al, 1999; Yung et al., 1996). Psychometric properties for both instruments have been extensively studied, and predictive validity for these instruments has been excellent for the conversion to psychosis outcome (AUC=0.85) (Oliver et al, 2022). Interrater reliability (IRR) for CHR-P ascertainment has also been excellent, both for the SIPS (median kappa across 16 published samples 0.89) (Woods et al, 2019) and the CAARMS (median across three studies 0.845) (Fusar-Poli et al, 2012; Miyakoshi et al, 2009; Paterlini et al, 2019). IRR for attenuated positive symptoms has also been excellent for the SIPS (median ICC across 21 published samples 0.88) (Woods et al., 2019) and CAARMS (median ICC or Pearson *r* across eight studies 0.89) (Braham et al, 2014; Fusar-Poli et al., 2012; Lho et al, 2021; Miyakoshi et al., 2009; Paterlini et al., 2019; Wang et al, 2022; Yokusoglu et al, 2021; Yung et al, 2005).

Recently the US National Institute of Mental Health (NIMH) has spearheaded an effort to harmonize these two instruments (Addington et al, 2023). Harmonization was needed despite identical attenuated positive symptom content and general overall similarity (Schultze-Lutter et al, 2013) because of six important differences in: 1) organization of attenuated positive symptom content into items (Table 1), 2) scaling of items, 3) conceptualization of severity, 4) quantifying symptom frequency, 5) frank psychosis diagnosis criteria (Table 2), and 6) CHR-P syndrome criteria (Tables 3-5).

**Table 1.** Content Comparison across SIPS, CAARMS, and PSYCHS items

| PSYCHS Item | SIPS Item | CAARMS Item |
| --- | --- | --- |
| P1 Unusual Thoughts and Experiences | P1 Unusual Thought Content | P1 Unusual Thought Content |
| P2 Suspiciousness/Paranoia | P2 Suspiciousness | P2 Non-Bizarre Ideas |
| P3 Unusual Somatic Ideas | P1 Unusual Thought Content | P2 Non-Bizarre Ideas |
| P4 Ideas of Guilt | P1 Unusual Thought Content | P2 Non-Bizarre Ideas |
| P5 Jealous Ideas | P1 Unusual Thought Content | P2 Non-Bizarre Ideas |
| P6 Unusual Religious Ideas | P1 Unusual Thought Content | P2 Non-Bizarre Ideas |
| P7 Erotomantic Ideas | P3 Grandiose Ideas | P2 Non-Bizarre Ideas |
| P8 Grandiosity | P3 Grandiose Ideas | P2 Non-Bizarre Ideas |
| P9 Auditory Perceptual Abnormalities | P4 Perceptual Abnormalities | P3 Perceptual Abnormalities |
| P10 Visual Perceptual Abnormalities | P4 Perceptual Abnormalities | P3 Perceptual Abnormalities |
| P11 Olfactory Perceptual Abnormalities | P4 Perceptual Abnormalities | P3 Perceptual Abnormalities |
| P12 Gustatory Perceptual Abnormalities | P4 Perceptual Abnormalities | P3 Perceptual Abnormalities |
| P13 Tactile Perceptual Abnormalities | P4 Perceptual Abnormalities | P3 Perceptual Abnormalities |
| P14 Somatic Perceptual Abnormalities | P4 Perceptual Abnormalities | P3 Perceptual Abnormalities |
| P15 Disorganized Communication | P5 Disorganized Communication | P4 Disorganised Speech |

Green text indicates the same health experience content is contained in the same item in SIPS 5.6.1 and CAARMS 2015; red text indicates the same health experience content is contained in different items in SIPS 5.6.1 and CAARMS 2015.

**Table 2.** Frank psychosis criteria for the SIPS and the CAARMS and the harmonized PSYCHS criteria

| Instrument | Psychosis Criteria |  |  |
| --- | --- | --- | --- |
|  | SIPS 5.6.1 | CAARMS 2015 | PSYCHS |
| Severity | Any of SIPS P1-P5=6 | Any of CAARMS P1-P4=6, or P3=5 | Any of PSYCHS P1-P15=6 |
| Timeframe | Lifetime | Lifetime | Lifetime |
| Frequency | One hour per day or more at an average frequency of four days a week | 3 or more days a week - one hour or more a day, or at least daily | 3 or more days a week - one hour or more a day, or at least daily |
| Duration | One month | One week or longer unless new or increased antipsychotic | One week or longer unless new or increased antipsychotic |
| Danger | Frequency and duration waived if seriously disorganizing or dangerous | None | Frequency and duration waived if imminently dangerous* |

\* physically or to personal dignity or to social/family networks
Red text indicates differences between SIPS and CAARMS, green text indicated harmonized criteria for psychosis

**Table 3.**
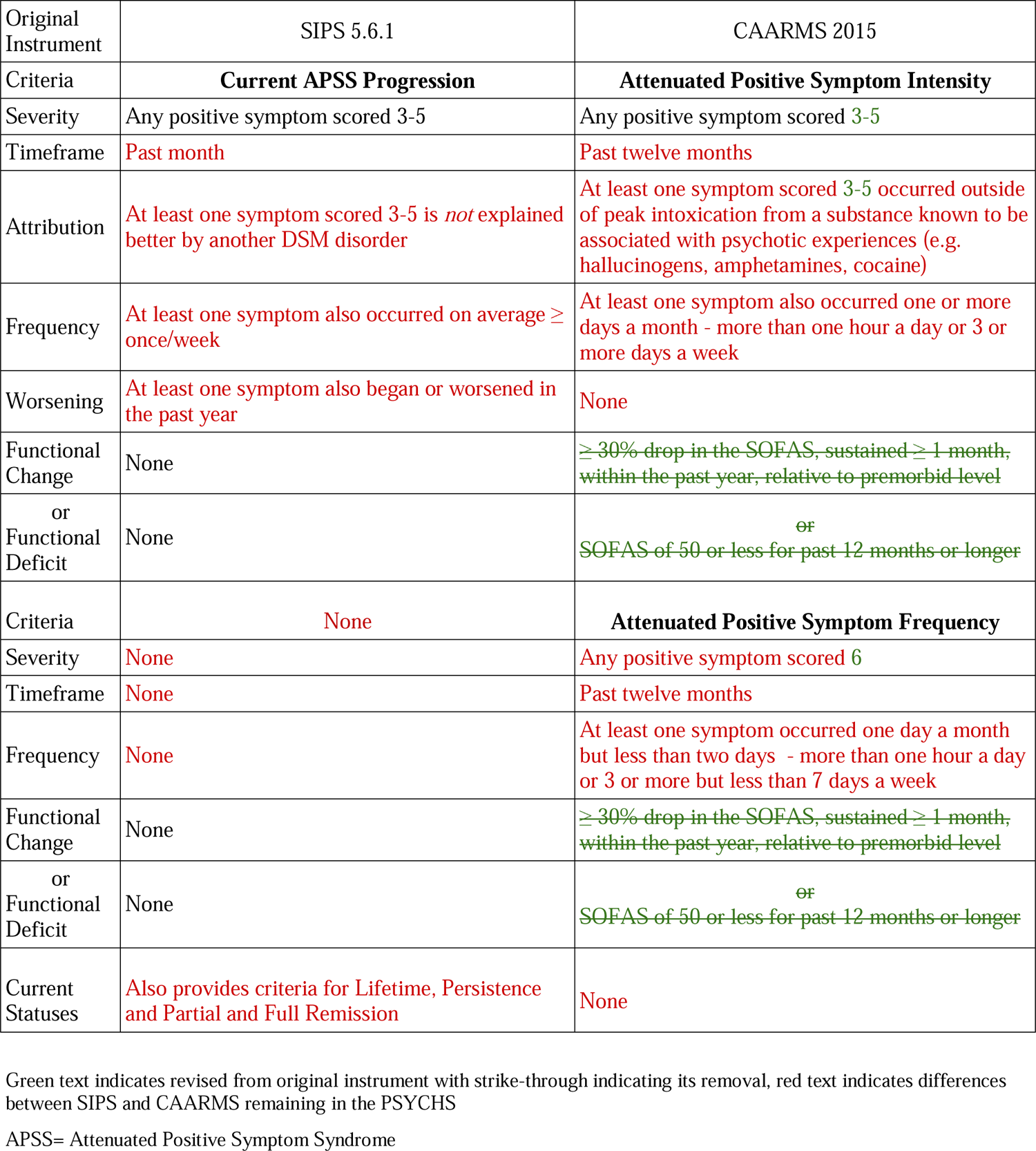
PSYCHS CHR-P criteria based on attenuated positive symptoms

These six differences make it challenging if not impossible to translate severity scores or diagnoses from one instrument to another and consequently generate uncertainty about comparing findings from studies that use one but not the other (Addington et al., 2023). In fact, some authors have described the state of assessment in the CHR-P field as one of “near-Babylonian” confusion (Schultze-Lutter et al, 2011). Using both instruments in a single study has generally been impractical due to participant burden and cost considerations. Therefore harmonization seemed to be the only solution.

The goal of this effort was to create a new instrument that harmonizes the CAARMS and the SIPS to the degree feasible based on current knowledge. The harmonized instrument is called <u>P</u>ositive <u>SY</u>mptoms and Diagnostic Criteria for the <u>C</u>AARMS <u>H</u>armonized with the <u>S</u>IPS (PSYCHS). It generates fully harmonized positive symptom ratings, provides for scoring of all CAARMS and SIPS positive symptom items from a single interview, fully harmonizes psychosis criteria, and generates partially harmonized CHR/UHR diagnostic criteria for both the CAARMS and the SIPS. This paper describes the methods and results for the harmonization in detail, including limits to harmonization; it also briefly outlines our implementation in the ongoing Accelerating Medicines Partnership® Schizophrenia (AMP® SCZ) observational study (Brady et al, 2023).

## 2 METHODS

### 2.1 Harmonization process

The initial harmonization process began when the NIMH hosted a workshop on February 13^th^ and 14^th^ 2020, attended by 38 international participants and described in the companion report (Addington et al., 2023). After the workshop, the lead experts for the SIPS and CAARMS (SWW and ARY) began a series of videoconference meetings in April 2020 facilitated by a NIMH program officer (SAW). These meetings considered workshop recommendations and unresolved issues and were generally held weekly for two hours. Beginning in January 2021, additional members with extensive practical experience with the CAARMS (SP, MJK) and the SIPS (BCW) joined these meetings.

Meeting time was spent reviewing the literature, comparing item content between SIPS version 5.6.1 (Keefe et al, 2021; Walsh, 2021) and CAARMS 2015 (Yung et al, 2015), ensuring that all attenuated positive symptom content in both instruments was captured in the PSYCHS by verbatim interviewer inquiries, reformulating the joint item content into new and distinct items (Table 1), ensuring the consistency of measurement concepts across items, harmonizing scaling, ensuring that the harmonized scale anchors for each item were distinct, ordered, and graded according to similar intervals within each measurement concept, and crafting interviewer and scoring instructions. All decisions were made by consensus, and minutes were taken by SAW.

### 2.2 Limits to harmonization

The initial charge in the NIMH-hosted workshop was to fully harmonize the two instruments. The workshop ended with incomplete progress, however, due to the number and difficulty of the challenges presented. After more than a year of intensive weekly meetings, the working group members agreed that it was possible to fully harmonize the assessment of attenuated positive symptoms. It was also possible to fully harmonize the diagnostic criteria for frank psychosis used for excluding CHR-P at ascertainment and for determining conversion/transition to frank psychosis. Although some progress was made in harmonizing CHR-P syndrome criteria, in the end, the different conceptualizations of the CHR-P syndrome proved too difficult to reconcile, and the group focused on designing the PSYCHS to generate data for both CAARMS and SIPS CHR-P syndrome criteria.

PSYCHS developers intended to keep the average administration time for the initial assessment version to no more than 90 minutes on average and no more than 60 minutes on average for the follow-up version, both broadly consistent with CAARMS and SIPS administration times. To meet these participant- and interviewer-burden goals, it was necessary to focus exclusively on diagnostic assessment and on attenuated positive symptoms that are required for that assessment. As a result, assessments for negative, disorganized, and general symptoms in the SIPS and for cognitive change, negative symptoms, behavioral change, motor/physical changes, and general psychopathology in the CAARMS were not included.

### 2.3 Implementation process

Harmonization was completed by December 2021. Work then shifted to implementing the instrument in Research Electronic Data Capture (REDCap) and the in-house Research Project Management System (RPMS), in collaboration with three projects included in the AMP SCZ consortium: the Psychosis-risk Outcomes Network (ProNET; SWW, CEB, and JMK, PIs), the Trajectories and Predictors in the CHR for Psychosis Population: Prediction Scientific Global Consortium (PRESCIENT; BN and PJM, PIs), and the Psychosis Risk Evaluation, Data Integration and Computational Technologies (PREDICT) Data Processing, Analysis and Coordination Center (DPACC; MES and RSK, PIs).

Implementation of the initial assessment version in REDCap and RPMS was completed by May 2022. Rater training and certification then began, for which JA and AN joined the working group meetings, and consensus calls were organized. Data collection for the initial assessment version began in the large observational study component of AMP SCZ in June 2022. Implementation of the follow-up version was completed by July 2022.

## 3 RESULTS

Results are presented for the fully harmonized acquisition of attenuated positive symptoms, the fully harmonized psychosis determination, and the partially harmonized and parallel SIPS/CAARMS CHR/UHR determinations. Materials available and current use in AMP SCZ are also briefly described.

### 3.1 Fully harmonized attenuated positive symptom acquisition

Full harmonization of the CAARMS and the SIPS attenuated positive symptoms was achieved in the areas of: symptom content, content organization into items, measurement concepts within each item, scaling of severity level, anchors for each level for each measurement concept for each item, fully-structured inquiries about patient health experiences mapping onto each item, and scoring of severity.

Figure 1 shows the conceptual framework underlying attenuated positive symptom acquisition in the PSYCHS. Following US Food and Drug Administration guidance (U.S. Department of Health and Human Services et al, 2022), the framework consists of a conceptual model and a measurement model. In the conceptual model, attenuated positive symptom-related health experiences resulting from CHR-P are organized into 15 distinct symptoms. Each of these is captured in the PSYCHS by two or more verbatim Inquiries and semi-structured Follow-up Questions. These health experiences are organized into three general concepts: 1) attenuated delusions, 2) attenuated hallucinations, and 3) attenuated thought disorder. Together the three general concepts form the concept of interest (Overall Attenuated Positive Symptom Burden of the Clinical High Risk Syndrome for Psychosis). In the measurement model, the PSYCHS is a Clinical Outcomes Assessment (COA) instrument as defined by FDA (U.S. Department of Health and Human Services et al., 2022) and yields a CHR-P attenuated positive symptom severity index comprising severity scores from 15 measurement items corresponding to 15 health experience areas captured by the PSYCHS.

**Figure 1.**
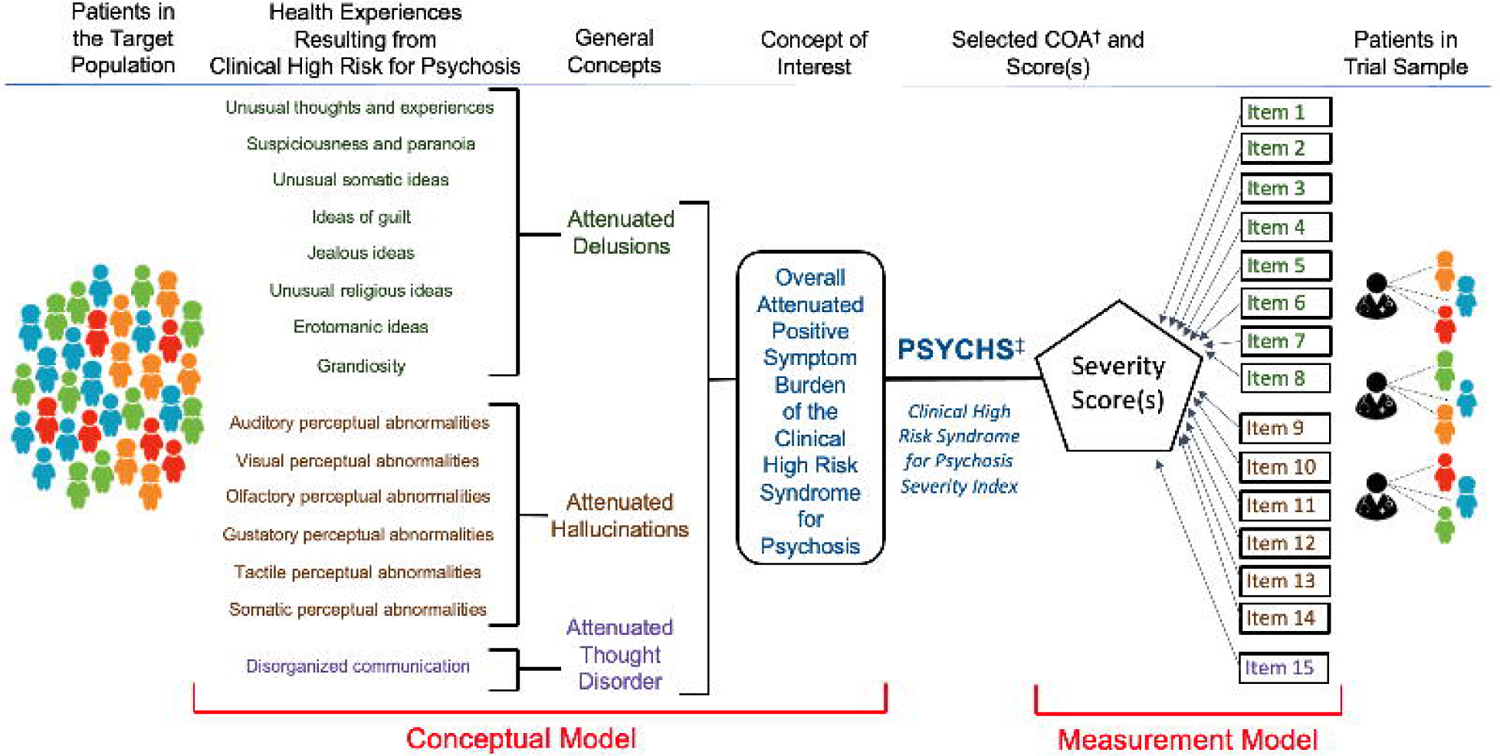
COA^†^ conceptual framework for the PSYCHS^‡^ symptom severity assessment. The conceptual framework consists of a conceptual model (left side of panel) and a measurement model (right side of panel). In the conceptual model, attenuated positive symptom-related health experiences resulting from the Clinical High Risk Syndrome for Psychosis are organized into 15 distinct symptoms. These health experiences are organized into three general concepts: 1) attenuated delusions, 2) attenuated hallucinations, and 3) attenuated thought disorder. Together the three general concepts form the concept of interest. In the measurement model, 15 measurement items corresponding to the health experience areas captured by the PSYCHS yield severity scores that in turn are used to compute a Clinical High Risk Syndrome for Psychosis severity index. † Clinical Outcomes Assessment, ‡ <u>P</u>ositive <u>SY</u>mptoms and Diagnostic Criteria for the <u>C</u>AARMS <u>H</u>armonized with the <u>S</u>IPS

#### 3.1.1 Content coverage

Review of the separate instrument instructions, manuals, and positive symptom inquiries and items revealed identical positive symptom content across the SIPS and CAARMS.

#### 3.1.2 Content organization into items

Although positive symptom content was identical, the same content was organized across the SIPS and the CAARMS into different items and into a different number of items based on differing formulations of psychopathology. Table 1 shows how attenuated positive symptom content mapped across the instruments. For example, unusual somatic ideas were captured in P1 of the SIPS (Unusual Thought Content) because they were neither paranoid nor grandiose in nature and so did not belong in SIPS P2 or P3; the CAARMS, however, captured unusual somatic ideas in P2 (Non-Bizarre Ideas) because they were not bizarre in the sense that they were theoretically possible. Another example is grandiosity, which was considered an independent item in the SIPS (P3) but designated as a component of Non-Bizarre Ideas (P2) in the CAARMS. No procedure could be devised to harmonize the two instruments by reorganizing content into just a handful of items without losing the integrity of individual items that have been strongly predictive of future psychosis in previous studies (Cannon et al, 2016). Thus Unusual Somatic Ideas, Ideas of Guilt, Jealous Ideas, and Unusual Religious Ideas each required separate items in the PSYCHS (Table 1 and Figure 1).

Since at least nine items would be needed to capture all of the CAARMS and SIPS attenuated positive symptom content, consideration was given to whether further splitting was desirable. Erotomania was separated from other forms of grandiosity, consistent with evidence that erotomania can constitute a distinct psychotic syndrome (Segal, 1989). Previously erotomania was rated in the SIPS under P3 grandiosity and in the CAARMS under P2 Non-Bizarre Ideas. We elected to divide the single perceptual abnormalities in both CAARMS and SIPS into six items: auditory, visual, olfactory, gustatory, tactile, and somatic, based on evidence that the combined perceptual abnormalities items predicted future psychosis poorly (Katsura et al, 2014; Perkins et al, 2015; Zhang et al, 2018) and mixed evidence that abnormalities of specific perceptual modalities may predict future psychosis differently (Ciarleglio et al, 2019; Lehembre-Shiah et al, 2017; Niles et al, 2019). Content of Disorganized Communication Expression was already harmonized (PSYCHS P15). Thus the PSYCHS was formulated with 15 attenuated positive symptom items (Table 1).

One experience, nihilistic ideas, had been captured in the CAARMS under P2 Non-Bizarre Ideas and in the SIPS under P1 Unusual Thought Content. We considered formulating nihilistic ideas into a separate item, perhaps along with perplexity and delusional mood, but in the end felt that additional psychopathology research was needed to properly construct a severity gradient and that for now nihilism should be placed within PSYCHS P1 (Unusual Thoughts and Experiences).

The name for P1 in both SIPS and CAARMS is Unusual Thought Content, and both instruments organize mental events and experiences such as thought insertion into this item. This organization is consistent with psychopathological classification of thought insertion as a delusion rather than a hallucination (American Psychiatric Association, 2013) due to the lack of a sensory component. Following Fish (Hamilton, 1984), who considered mental events such as thought insertion to be *experiences,* the name for PSYCHS P1 was changed to Unusual Thoughts and Experiences.

#### 3.1.3 Attenuated positive symptom measurement concepts

Positive symptom severity is complex and multidimensional, and symptom severity anchors in both SIPS and CAARMS have always contained mixtures of measurement concepts in the item anchors. Attention to distinguishing measurement concepts within the anchors has become more detailed and explicit with subsequent revisions for each instrument. With the revision from version 5.6 to 5.6.1 in 2017, SIPS anchors have been designed so that each item contains a graded description of each measurement concept for each severity level.

This structure was maintained in the PSYCHS. Each item is conceptualized as composed of, and each scale level for each symptom is closely anchored for, three or four measurement concepts: 1) symptom description (all items); 2) symptom tenacity (for attenuated delusion items P1 to P8), symptom source (for attenuated hallucination items P9 to P14), or symptom self-correction (for attenuated disorganized communication item P15); 3) distress due to the symptom (all items except P8 Grandiosity); and 4) interference (with other thoughts, feelings, social relations and/or behavior) due to the symptom (all items).

The measurement concepts are synthesized into a single rating for the item as follows: the first two measurement concepts are co-primary and generally determine the item’s single rating. For example, if an interviewer judges that symptom description matches anchor text for 5, and symptom tenacity/source/self-correction also matches anchor text for 5, the item single rating for that timeframe is 5.

The third and fourth measurement concepts (distress and interference) are secondary. In the example above, the secondary measurement concepts do not contribute to the single rating. The secondary measurement concepts only contribute to the single rating in the situation when the interviewer determines that the co-primary measurement concepts do not agree. For example, when the interviewer judges that symptom description matches anchor text for 4 but symptom tenacity/source/self-correction matches anchor text for 5, or vice-versa, the interviewer should take into account anchor text for distress due to the symptom and for interference due to the symptom. If *either* distress *or* impairment due to the symptom matches anchor text in the 5 or 6 range, the single rating for that item will be 5. If *both* distress *and* impairment due to the symptom match anchor text in the 4 or lower range, the single rating for that item will be 4.

Among the attenuated hallucinations items (P9-P14), the focus of the secondary measurement concept is on the perceived source of the perception, in other words the degree to which the experience is perceived to arise from a real source as opposed to arising from one’s own thoughts. The concept of perceived source is derived from the CAARMS and represents a change for the SIPS. Previously the SIPS P4 Perceptual Abnormalities item considered the degree to which the sensory experience was believed to be real instead of the degree to which it was perceived as real. Colleagues occasionally pointed out the inconsistency in the SIPS in having a perceptual item rely on a delusional interpretation, and so the SIPS developers on the team were amenable to adopt the CAARMS procedure. Independent perceptual and delusional items may facilitate research focusing on the co-occurrence and sequencing of onset of attenuated delusions and hallucinations (Mourgues et al, 2023; Smeets et al, 2015).

Thus the PSYCHS gives strong and often exclusive priority to the two primary measurement concepts in determining severity/intensity. The rationale for this approach was that distress or disability associated with attenuated positive symptoms may be affected by other factors in addition to actual attenuated positive symptom severity, such as depression or anxiety, consistent with a recent empirical analysis (Wilson et al, 2020).

#### 3.1.4 Harmonized attenuated positive symptom item scaling

Attenuated positive symptom item scaling differed between the CAARMS and SIPS. For the SIPS, the fully psychotic range was limited to level 6, the subsyndromal or CHR range was 3-5, and the non-pathological range was 0-2 for all five attenuated positive symptom items. For the CAARMS, the same was true for items P1 and P2, but for CAARMS P3 (perceptual abnormalities) the fully psychotic range was 5-6, the subsyndromal or CHR range was 3-4, and the non-pathological range was 0-2, while for CAARMS P4 (conceptual disorganization), the fully psychotic range was limited to level 6, the subsyndromal or CHR range was 4-5, and the non-pathological range was 0-3.

As part of the harmonization process, CAARMS developers felt that consistency across items was an advantage for raters, and anchor content for the PSYCHS was crafted so that the severity gradient reflected frank psychosis at level 6, the subsyndromal or CHR range at 3-5, and the non-pathological range at 0-2 for all 15 attenuated positive symptom items. On careful inspection of the original instrument anchors, it was possible to meld content from the two instruments so that, for example, level 6 on the PSYCHS attenuated hallucinations items retained consistency with levels 5 and 6 from CAARMS P3 while also retaining consistency with the distinction between levels 5 and 6 on SIPS P4.

The labels for the anchor levels also differed slightly across the original instruments. For SIPS 5.6.1, levels 0-6 were labeled, respectively: Absent; Questionably Present; Mild; Moderate; Moderately Severe; Severe but not Psychotic; and Severe and Psychotic. For CAARMS 2015, levels 0-6 were labeled, respectively: Never, absent; Questionable; Mild; Moderate; Moderately severe; Severe; and Psychotic & severe. The working group agreed that these could be fully harmonized as: Absent; Questionable; Mild; Moderate; Marked; Severe but not Psychotic; and Psychotic and Very Severe.

#### 3.1.5 Harmonized attenuated positive symptom item anchors

Once the scaling challenges were surmounted, it was conceptually straightforward to meld text from the original instrument anchors into harmonized text for each measurement concept, for each anchor, and for each item. Careful attention was paid so that within each measurement concept for each item, the seven (0-6) levels described different severity levels of the same content, that the seven levels were each distinct from one another, that each adjacent level was ordered relative to its neighbors, and that a consistent increasing gradient of severity existed across levels within each measurement concept. The anchor sets for each item were further scrutinized for consistency with the anchor labels, such that, for example, the word “marked” was not used in an anchor under Severe but not Psychotic. Lastly, the anchors within each measurement concept were evaluated across items, so that, for example, the same words were not used for differing levels across items.

#### 3.1.6 Harmonized attenuated positive symptom inquiries

Since the CAARMS and the SIPS covered identical overall positive symptom content, harmonizing verbatim inquiries about participant’s health experiences was relatively straightforward. The two sets of inquiries were merged, and redundancies were eliminated.

#### 3.1.7 Concept of severity

The two instruments conceptualize severity similarly in most regards, as reviewed above, and when there is variability of severity within the measurement interval both instruments capture the highest severity during that interval. There is one important difference, however (Addington et al., 2023). The SIPS conceptualizes the synthesis of the measurement concepts for a particular item over the past month as *severity*. The CAARMS conceptualizes the same measurement concepts over the same recall interval as *intensity* rather than as severity and adds an additional severity measurement concept of symptom *frequency*. Intensity and frequency are then combined to yield CAARMS severity. Since this difference could not be harmonized, the PSYCHS generates ratings for both SIPS and CAARMS conceptualizations of severity. To acknowledge this difference, the synthesis of the four harmonized severity-relevant measurement concepts in the PSYCHS items (not including frequency) is termed *severity/intensity* within the instrument. In addition, a new severity score native to the PSYCHS is calculated as the sum of PSYCHS items P1-P15 (range 0-90).

#### 3.1.8 SIPS item generation and scoring of SIPS severity

For SIPS attenuated positive symptom severity, five items are generated from the PSYCHS, consistent with the mapping shown in Table 1. SIPS P1 severity is calculated as equal to the highest of PSYCHS items 1, 3, 4, 5, and 6 severity/intensity. SIPS P2 severity equals PSYCHS item 2. SIPS P3 severity is equal to the higher of PSYCHS items 7 and 8. SIPS P4 severity is equal to the highest of PSYCHS items 9-14. SIPS P5 severity equals PSYCHS item 15. The SIPS total attenuated positive symptom severity score is the sum of SIPS items P1-P5 (range 0-30) as per usual practice.

#### 3.1.9 CAARMS item generation and scoring of CAARMS severity

The PSYCHS incorporates the CAARMS 0-6 frequency ratings for each of the 15 items, with minor adjustments when rating the past month timeframe. Consistent with the mapping shown in Table 1 and with previous practice (Hartmann et al, 2020; Morrison et al, 2012), CAARMS P1 severity is equal to the product of PSYCHS P1 severity/intensity and frequency. CAARMS P2 severity is equal to the highest of the seven products of severity/intensity and frequency for PSYCHS items P2-P8. CAARMS P3 severity is equal to the highest of the six products of severity/intensity times frequency for PSYCHS items P9-14. The CAARMS total attenuated positive symptom severity score is the sum of the CAARMS P1-P4 severity/intensity-frequency products (range 0-144).

### 3.2 Fully harmonized psychosis determination

CAARMS 2015 and SIPS 5.6.1 criteria for frank psychosis, used for excluding fully psychotic participants at study ascertainment and as criteria for conversion/transition to psychosis during study follow-up, differed in four of five domains, being identical only on the rating time frame (Table 2). Since conversion/transition was a frequently used outcome measure, the authors felt that it was essential to harmonize these criteria. Moreover, a study wherein both SIPS and CAARMS criteria were derived from a single modified CAARMS interview found considerable disagreement on presence of frank psychosis (Fusar-Poli et al, 2016b). The harmonization of attenuated positive symptom severity (see above) permitted full agreement in the severity domain, and consensus was reached on the remaining three domains, as described in sections 3.2.1. and 3.2.2. The fully-harmonized psychosis criteria are included in Appendix S1.

#### 3.2.1 Harmonization of duration and frequency criteria for frank psychosis

The SIPS has required a duration of fully psychotic symptoms of one month to qualify for psychotic disorder, consistent with DSM-5 criteria for schizophrenia (American Psychiatric Association, 2022). CAARMS duration criteria were greater than or equal to one week. In practice the SIPS duration and frequency criteria could permit a frank psychosis determination in as little as 16 days if the psychotic-level symptoms were experienced daily (which averages to four days a week for a month). However, practitioners and patients and their families were often reluctant to wait that long to institute treatment for frank psychosis, and the SIPS developers were agreeable to adopt the CAARMS frequency and duration criteria (Table 2).

#### 3.2.2 Harmonization of the frank psychosis dangerousness criterion

The SIPS waiver of frequency and duration criteria when fully psychotic symptoms were disorganizing or dangerous had been a sticking point in the initial NIMH workshop (Addington et al., 2023). This waiver was meant in part to mitigate the risk of delayed SIPS diagnosis of psychosis due to the one month duration criterion when the need for treatment was immediate. The shorter CAARMS duration requirement, and its exception for cases that received new or increased antipsychotic medication, mitigated the risks associated with the longer SIPS duration criteria to some extent. However, those risks were not mitigated entirely. In addition, the SIPS waiver of the frequency and duration criteria when fully psychotic symptoms were disorganizing or dangerous also functioned to mitigate a difficulty with the duration criteria when evaluating a person shortly after onset and when frank psychosis was clear-cut. This difficulty is that clinicians and researchers can be left in limbo without a psychosis determination if the participant is unable to be reevaluated a week later. That situation can occur around the time of conversion/transition if frank psychosis leads the participant to disengage from a clinical service or to be unable or unwilling to continue research participation. The SIPS waiver of the frequency and duration criteria resolves this difficulty in cases where symptoms are so clearly indicative of frank psychosis that they are associated with danger to self or others.

During the course of the intensive follow-up meetings, the CAARMS developers found these arguments reasonably compelling and were agreeable to adopt the SIPS waiver, so long as the phrase “seriously disorganizing or dangerous” was reworded. SIPS developers had on occasion been asked questions about what “seriously disorganizing” meant, or needed to correct confusion between “disorganizing” and disorganization symptoms, and thus the authors agreed on substituting “imminently dangerous, physically or to personal dignity or to social/family networks.” These criteria enable a psychosis diagnosis to be made at a single visit when, for example, a person’s dignity and reputation are threatened by psychotic behavior or when their or another’s life is endangered due to psychotic thinking or behavior.

### 3.3 Partially harmonized and parallel CHR/UHR determination

Following the CAARMS, the SIPS has always generated three CHR/UHR syndromes based on the same three principles: 1) presence of attenuated positive symptoms (CAARMS Attenuated Positive Symptom Intensity and Attenuated Positive Symptom Frequency/SIPS Attenuated Positive Symptoms Syndrome, Table 3), 2) presence of brief fully psychotic symptoms (CAARMS Brief Limited Intermittent Psychotic Symptoms/SIPS Brief Intermittent Psychosis Syndrome, Table 4), and 3) presence of trait vulnerability and functional decline (CAARMS Vulnerability group/SIPS Genetic Risk and Functional Deterioration, Table 5). The detailed definitions for each of the three CHR/UHR syndromes differed, however. In the end the working group was able to reconcile these differences only to a relatively minor degree (sections 3.3.1 and 3.3.2).

**Table 4.** PSYCHS CHR-P criteria based on brief fully psychotic symptoms

| Original Instrument | SIPS 5.6.1 | CAARMS 2015 |
| --- | --- | --- |
| Criteria | **Current BIPS Progression** | **Brief Limited Intermittent Psychotic Symptoms (BLIPS)** |
| Severity | Any positive symptom scored 6 | Any positive symptom scored 6 |
| Timeframe | Past month | Past twelve months |
| Attribution | At least one symptom scored 6 is *not* explained better by another DSM disorder | At least one symptom scored 6 occurred outside of peak intoxication from a substance known to be associated with psychotic experiences (e.g. hallucinogens, amphetamines, cocaine) |
| Frequency | At least one symptom also occurred ≥ several minutes a day at least once in the past month | At least one symptom also occurred three or more days a week - more than one hour a day or at least daily |
| Duration | None | Less than one week |
| Worsening | At least one symptom also began or worsened in the past three months | None |
| Functional Change | None | ~~$\geq 30\%$ drop in the SOFAS, sustained $\geq 1$ month, within the past year, relative to premorbid level~~ |
| or Functional Deficit | None | ~~or SOFAS of 50 or less for past 12 months or longer~~ |
| Current Statuses | Also has criteria for Lifetime, Persistence and Partial and Full Remission | None |

Green text indicates revised from original instrument with strike-through indicating its removal, red text indicates differences between SIPS and CAARMS remaining in the PSYCHS
BIPS=Brief Intermittent Psychosis Syndrome
BLIPS=Brief Limited Intermittent Psychotic Symptoms

**Table 5.**
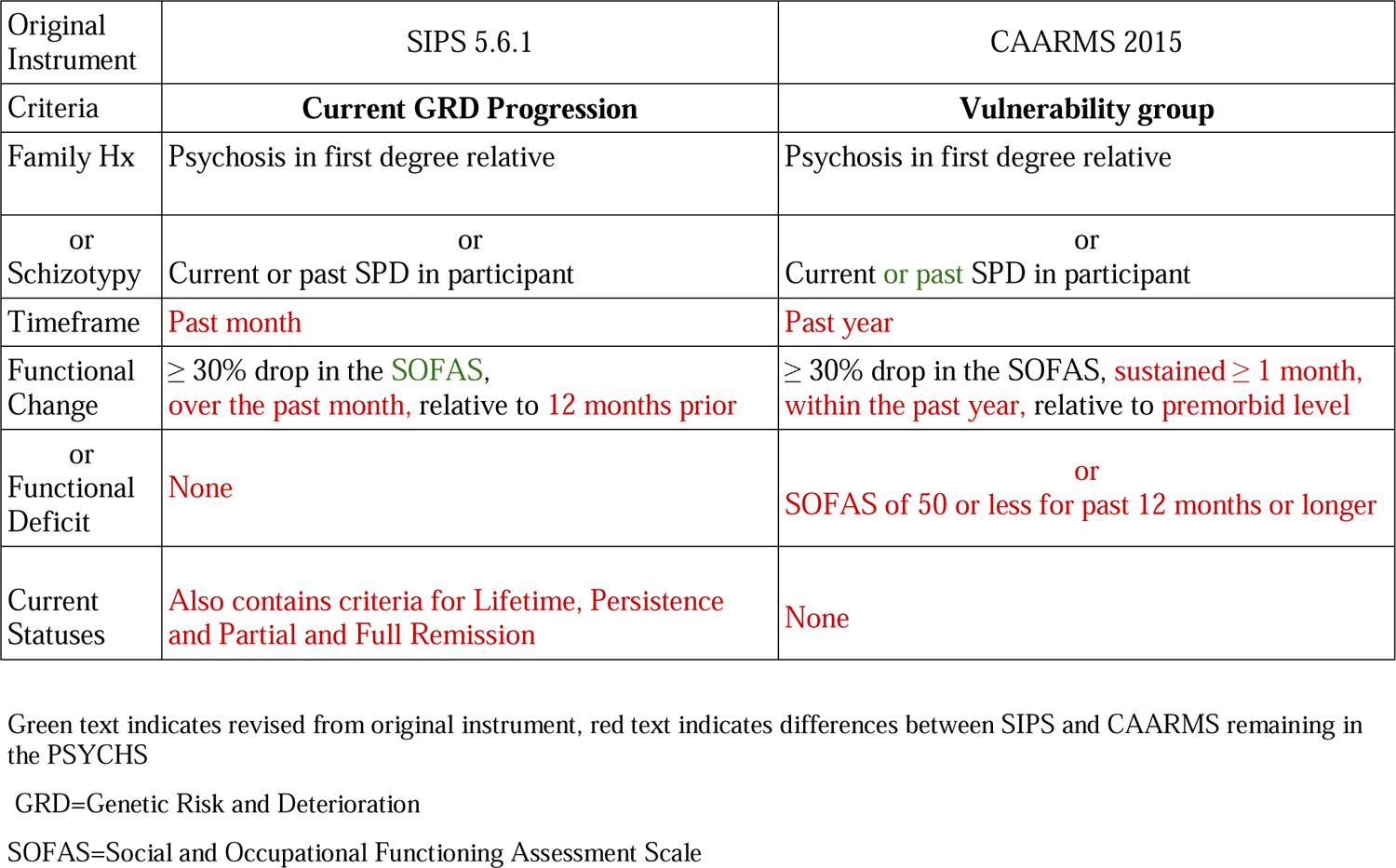
PSYCHS CHR-P criteria based on trait vulnerability and functional impairment

| Original Instrument | SIPS 5.6.1 | CAARMS 2015 |
| --- | --- | --- |
| Criteria | **Current GRD Progression** | **Vulnerability group** |
| Family Hx | Psychosis in first degree relative | Psychosis in first degree relative |
| or<br>Schizotypy | or<br>Current or past SPD in participant | or<br>Current **or** past SPD in participant |
| Timeframe | **Past month** | **Past year** |
| Functional Change | ≥ 30% drop in the **SOFAS**,<br>**over the past month**, relative to **12 months prior** | ≥ 30% drop in the SOFAS, **sustained ≥ 1 month**,<br>**within the past year**, relative to **premorbid level** |
| or<br>Functional Deficit | **None** | **or**<br>**SOFAS of 50 or less for past 12 months or longer** |
| Current Statuses | **Also contains criteria for Lifetime, Persistence and Partial and Full Remission** | **None** |

Green text indicates revised from original instrument, red text indicates differences between SIPS and CAARMS remaining in the PSYCHS
GRD=Genetic Risk and Deterioration
SOFAS=Social and Occupational Functioning Assessment Scale

For the syndromes based on presence of attenuated positive symptoms (Table 3), the achievement of symptom severity harmonization offered promise, and the frequency criteria could potentially have been harmonized, but neither investigator group could compromise on the several remaining differences. The SIPS required attenuated positive symptoms to have been present in the past month and considered them in remission if they were no longer present in the past month (Woods et al, 2014), while the CAARMS permitted attenuated positive symptoms to have been present at any time in the past year. A compromise period of six months was proposed at the workshop (Addington et al., 2023), but during the extended discussions SIPS developers could not agree that symptoms no longer present in the past month should not be considered in at least partial remission. Moreover, the SIPS requires one or more attenuated positive symptoms to have begun or worsened in the past year, while the CAARMS does not. SIPS developers considered that epidemiologic (Schultze-Lutter et al, 2014) and other (Addington et al., 2023; Brucato et al, 2019; Woods et al., 2014) evidence suggested that the worsening criterion favorably excluded large numbers of patients who were no longer at high risk of conversion/transition, while CAARMS developers considered that the SIPS unfavorably excluded large numbers of patients with a need for treatment.

Lastly, the SIPS developers preferred accordance with the DSM-5 principle of parsimony such that a second diagnosis is not needed if all of its features are accounted for by another disorder, whereas the CAARMS was often employed on its own in a clinical context and so CAARMS developers were concerned that excluding patients from a CAARMS grouping could cause them to be excluded from care. Unable to agree, the authors settled for requiring the PSYCHS to include questions that would generate both sets of CHR/UHR criteria.

The issues preventing full harmonization for the syndromes based on presence of brief fully psychotic symptoms (Table 4) were similar, as were the issues preventing full harmonization for the syndromes based on trait vulnerability and functional decline (Table 5).

#### 3.3.1 Modifications to CAARMS UHR criteria

CAARMS developers agreed to remove functioning criteria based on the Social and Occupational Functioning Assessment Scale (SOFAS) (Morosini et al, 2000) from the symptom-based UHR syndromes (Tables 3 and 4), harmonizing with the SIPS. These criteria were removed as it was acknowledged that (1) treatment of UHR individuals may be needed in the absence of functional decline and (2) removing the functional decline criterion would enable early intervention to prevent deterioration. At the initial NIMH workshop (Addington et al., 2023), the consensus had been that the field should abandon the CAARMS Vulnerability group/SIPS Genetic Risk and Deterioration subtype due to evidence that it was infrequent, especially in the absence of other subtypes, and did not predict onset of psychosis (Fusar-Poli et al, 2016a). AMP SCZ investigators, however, saw value in the subtype for the study of functional outcomes, leading to its retention. CAARMS developers agreed to base the Vulnerability group criteria on current or past schizotypal personality disorder (SPD) (First, 2014) rather than solely on current SPD (Table 5) after reviewing evidence that the diagnostic stability of SPD is not fully trait-like (Grilo et al, 2004). The modified CAARMS UHR criteria are included in Appendix S1.

#### 3.3.2 Modifications to SIPS CHR criteria

The SIPS has based the functional assessment requirement for Genetic Risk and Deterioration (GRD, Table 5) on the Global Assessment of Functioning (GAF) (Hall, 1995). Because of observations that GAF assessment of functioning was confounded by symptom severity (American Psychiatric Association, 1994), SIPS developers agreed to replace the GAF with the SOFAS, thus harmonizing the functional assessment scale with the CAARMS Vulnerability grouping. The modified SIPS GRD criteria are included in Appendix S1.

### 3.4 Available materials

The Interviewer Manual, training and certification materials, the Screening Instrument for ascertainment and initial severity rating, and the Follow-Up Instrument for serial rating of severity, conversion/transition, and remission, are freely available for use by the research community and will become accessible on the AMP SCZ website, developed by the PREDICT DPACC in collaboration with members of ProNET and PRESCIENT with input from NIMH staff: https://www.ampscz.org. Data sharing is otherwise not applicable to this article as no datasets were generated or analyzed for the current article.

The PSYCHS will be available in an on-line REDCap version and as a printable paper copy. The on-line version adaptively skips questions made unnecessary by previous interviewer entries, provides just-in-time guidance only when needed, and automatically conducts calculations for determining psychosis and CHR/UHR criteria. Information required at follow-up to determine new onset of psychosis or CHR-P syndromes is pulled automatically from previous visits. The coding of the calculations and branching logic for the PSYCHS in REDCap was carried out by members of PREDICT DPACC and ProNET, with testing across ProNET and PRESCIENT.

### 3.5 Current use

The PSYCHS is currently in use in the 42-site AMP SCZ (Brady et al., 2023) observational study (https://www.ampscz.org). As of December 2022, more than 100 interviewers had been trained and certified, more than 100 participants had undergone assessment with the Screening Instrument, and five coordinated weekly consensus calls were ongoing. All persons gave their informed consent prior to their inclusion in the study.

## 4 DISCUSSION

The principal finding of the present report is that it has been possible to harmonize the two most widely-used instruments for diagnosis and severity rating in individuals at clinical high risk for psychosis into one instrument, the PSYCHS. Full harmonization was achieved for attenuated positive symptom ratings and for psychosis diagnostic criteria, and the instrument generates partially harmonized CHR/UHR diagnostic criteria for both CAARMS and SIPS as well as severity scores for both CAARMS and SIPS.

The PSYCHS can be used instead of individual SIPS or CAARMS assessment for CHR-P ascertainment and attenuated positive symptom severity rating. When used in this way, future studies ideally would permit inclusion of participants who meet criteria for either CAARMS UHR or SIPS CHR Progression, and sensitivity analyses in a data supplement could then report whether findings differed by CAARMS vs SIPS ascertainment or when employing CAARMS vs SIPS severity ratings. This practice would be helpful in comparing findings across studies and with meta-analysis.

### 4.1 Strengths

The primary strengths of the PSYCHS are: 1) it harmonizes two instruments which both possess excellent psychometric properties, 2) the harmonization was conducted with great care by experts in both instruments, and 3) the attenuated positive symptom anchors provide detailed guidance for each of the 15 attenuated positive symptoms and are harmonized with particular attention to ensuring that anchors for each item are distinct, ordered, and graded according to similar intervals within each measurement concept. These changes are expected to yield even higher interrater reliability than already achieved with the original instruments and therefore improved signal detection. The on-line versions adaptively minimize administration time, missing data, and arithmetic errors.

### 4.2 Limitations

There are also a number of limitations to the PSYCHS in its current stage of development. First and foremost is the inability of the authors to fully harmonize the CHR-P diagnostic criteria. One of the difficulties is due to the limited evidence base available to contribute to deliberations. We are aware of only one study that reports conducting independent CAARMS and SIPS interviews in the same CHR participants (Kwon et al, 2012), and the report does not present diagnostic agreement or comparative predictive validity analyses. A recent study in relatives of patients with schizophrenia, however, also conducted independent interviews and reported 93% agreement, but agreement was largely due to the low prevalence of CHR in the sample of relatives. Of 17 cases diagnosed as CHR by either interview, the two interviews agreed on only 5 (29%) (Wang et al., 2022). Methods that rely on conducting only one interview and then estimating whether participants meet criteria for the other interview, while understandable in terms of limiting participant burden, may not be able to capture the other interview’s assessment accurately, given the differences in the details of the data collection required. Use of the PSYCHS in the AMP SCZ sample should enable analyses of diagnostic agreement between, and comparative predictive validity of, SIPS and CAARMS CHR criteria in a large sample of the same subjects. Based on these data it may be possible to fully harmonize the CAARMS and the SIPS CHR criteria in the future.

A second limitation derives from the PSYCHS being a new instrument whose psychometric properties need to be established. While interrater reliability has been excellent (section 1) for both the CAARMS and the SIPS, similarly excellent inter-rater reliability for the harmonized PSYCHS cannot be assumed. We will conduct reliability studies as part of the AMP SCZ observational study, as well criterion validity (Sheehan et al, 1998) and other psychometric studies, in accordance with guidelines from the US Food and Drug Administration (U.S. Department of Health and Human Services et al., 2022).

A third limitation is the synthesis of a single severity rating for each item across up to four measurement concepts. While the single severity rating has always been used for the SIPS and the CAARMS and could be considered a strength for assessment of outcomes, independent rating of each measurement concept may provide sufficient added value for the purposes of predicting outcome to offset the additional burden on participant and interviewer. For example, there is mixed evidence as to whether distress due to attenuated positive symptoms predicts future onset of frank psychosis independently from symptom severity (Nelson et al, 2022; Power et al, 2016; Pratt et al, in press; Rapado-Castro et al, 2015; Rekhi et al, 2019). We plan to investigate the independent rating of each measurement concept within AMP SCZ. Regarding the synthesis of measurement concepts by the interviewers, a cognitive debriefing study may be needed to demonstrate whether interviewers understand the method of synthesis.

Lastly, the PSYCHS contains 15 separate attenuated positive symptom items. While early experience in AMP SCZ indicates that administration times generally correspond to the intended 60-90 minutes, there have been exceptions, especially for an individual interviewer’s first case or two as they gain familiarity with navigating the instrument in REDCap or RPMS. Analyses from AMP SCZ will be used to determine whether certain items could be consolidated. The increased focus on positive symptoms also has required that negative, disorganized, and general symptoms must be rated using separate scales. When used to make CAARMS Vulnerability grouping/SIPS GRD determinations, the PSYCHS relies on the SOFAS (Morosini et al., 2000) for functional assessment, as well as on the Structured Clinical Interview for DSM-5 Personality Disorders (First, 2014) and the Family Interview for Genetics Studies (Maxwell, 1992) for determining presence of schizotypal personality disorder and first-degree family history of psychosis, respectively.

### 4.3 Summary

The <u>P</u>ositive <u>SY</u>mptoms and Diagnostic Criteria for the <u>C</u>AARMS <u>H</u>armonized with the <u>S</u>IPS semi-structured interview (PSYCHS) has been developed to harmonize the two most widely-used instruments for diagnosis and severity rating in patients at clinical high risk for psychosis (CHR-P). Use of the PSYCHS should facilitate comparing findings across studies in the CHR-P field.

## Data Availability

AMP SCZ data are held in the NIMH Data Archive and available at nda.nih.gov/ampscz.

## Acknowledgments

The Accelerating Medicines Partnership Schizophrenia (AMP SCZ) is a public-private partnership managed by the Foundation for the National Institutes of Health. The AMP SCZ research program is supported by contributions from the AMP SCZ public and private partners, including NIMH (U24MH124629, U01MH124631, and U01MH124639) and Wellcome (220664/Z/20/Z and 220664/A/20/Z). AMP SCZ Steering Committee members include: R. Benabou, L. Bilsland, L.S. Brady, T. Brister, F. Butlen-Ducuing, K. Duckworth, G.K. Farber, T.R. Farchione, B.A. Fischer, S. Frangou, S.T. Garcia, N. Gogtay, J.A. Gordon, R.K. Heinssen, W. Horton, B.R. Johnson, P.S. Joshi, C.A. Larrauri, S.H. Lisanby, G. Pandina, S.E. Roth, M. Sand, V.M. Sharma, B. Staglin, A. Travaglia, E. Velhorst, G. Wunderlich. Other AMP SCZ members not listed elsewhere include: M. Grabb, M. Hillefors, D. Janes, S.E. Morris, J. Pevsner. The views expressed in this paper are personal views of the authors and may not be understood or quoted as being made on behalf of or reflecting the position of the US Food and Drug Administration or the European Medicines Agency or one of its committees or working parties. The authors gratefully acknowledge the contributions of the following collaborators in the design and conduct of the AMP SCZ observational project: J.T. Baker, G. Cecchi, C. Gerber, M. Harms, L. Kambeitz-Ilankovic, M. Kubicki, S.-W. Kim, J.S. Kwon, A. Reichenberg, F.W. Sabb, Z. Tamayo, R. Upthegrove, S.K. Verma, I. Winter-van Rossum, D.H. Wolf, P. Wolff, S.J. Wood. All those acknowledged have given written permission.

## Data availability statement

AMP SCZ data are held in the NIMH Data Archive and available at nda.nih.gov/ampscz.

## References

Addington J, Stowkowy J, Liu L, Cadenhead KS, Cannon TD, Cornblatt BA, McGlashan TH, Perkins DO, Seidman LJ, Tsuang MT, Walker EF, Bearden CE, Mathalon DH, Santesteban-Echarri O, Woods SW (2019). Clinical and functional characteristics of youth at clinical high-risk for psychosis who do not transition to psychosis. Psychological Medicine 49(10), 1670–1677.

Addington J, Woods SW, Yung AR, Calkins ME, Fusar-Poli P (2023). Harmonizing the Structured Interview for Psychosis-Risk Syndromes (SIPS) and the Comprehensive Assessment of At-Risk Mental States (CAARMS): An initial approach. Early Interv Psychiatry in press.

American Psychiatric Association (1994). Diagnostic and Statistical Manual of Mental Disorders, Fourth Edition. American Psychiatric Association, Washington DC.

American Psychiatric Association (2013). Diagnostic and Statistical Manual of Mental Disorders, Fifth Edition. American Psychiatric Association, Arlington, VA.

American Psychiatric Association (2022). Diagnostic and Statistical Manual of Mental Disorders, Fifth Edition, Text Revision. American Psychiatric Publishing, Washington DC.

Andreou C, Bailey B, Borgwardt S (2019). Assessment and treatment of individuals at high risk for psychosis. BJPsych Advances 25(3), 177–184.

Brady LS, Laurrari CA, AMP SCZ Steering Committee (2023). Accelerating Medicines Partnership® Schizophrenia (AMP® SCZ): developing tools to enable early intervention in the psychosis risk state. World Psychiatry 22(1), 42–43.

Braham A, Bannour AS, Ben Romdhane A, Nelson B, Bougumiza I, Ben Nasr S, ElKissi Y, Ali BB (2014). Validation of the Arabic version of the Comprehensive Assessment of At Risk Mental States ( CAARMS) in Tunisian adolescents and young adults. Early Intervention in Psychiatry 8(2), 147–154.

Brucato G, First MB, Dishy GA, Samuel SS, Xu Q, Wall MM, Small SA, Masucci MD, Lieberman JA, Girgis RR (2019). Recency and intensification of positive symptoms enhance prediction of conversion to syndromal psychosis in clinical high-risk patients. Psychological Medicine, 1-9.

Cannon TD, Yu C, Addington J, Bearden CE, Cadenhead KS, Cornblatt BA, Heinssen R, Jeffries CD, Mathalon DH, McGlashan TH, Perkins DO, Seidman LJ, Tsuang MT, Walker EF, Woods SW, Kattan MW (2016). An individualized risk calculator for research in prodromal psychosis. American Journal of Psychiatry 173(10), 980–988.

Ciarleglio AJ, Brucato G, Masucci MD, Altschuler R, Colibazzi T, Corcoran CM, Crump FM, Horga G, Lehembre-Shiah E, Leong W, Schobel SA, Wall MM, Yang LH, Lieberman JA, Girgis RR (2019). A predictive model for conversion to psychosis in clinical high-risk patients. Psychological Medicine 49(7), 1128–1137.

Collins MA, Ji JL, Chung Y, Lympus CA, Afriyie-Agyemang Y, Addington JM, Goodyear BG, Bearden CE, Cadenhead KS, Mirzakhanian H, Tsuang MT, Cornblatt BA, Carrión RE, Keshavan M, Stone WS, Mathalon DH, Perkins DO, Walker EF, Woods SW, Powers AR, Anticevic A, Cannon TD (2022). Accelerated cortical thinning precedes and predicts conversion to psychosis: The NAPLS3 longitudinal study of youth at clinical high-risk. Molecular Psychiatry https://doi.org/10.1038/s41380-022-01870-7.

Daneault J-G, Stip E, Refer OSG (2013). Genealogy of instruments for prodrome evaluation of psychosis. Frontiers in Psychiatry 4, 25.

de Pablo GS, Besana F, Arienti V, Catalan A, Vaquerizo-Serrano J, Cabras A, Pereira J, Soardo L, Coronelli F, Kaur S, da Silva J, Oliver D, Petros N, Moreno C, Gonzalez-Pinto A, Diaz-Caneja CM, Shin JI, Politi P, Solmi M, Borgatti R, Mensi MM, Arango C, Correll CU, McGuire P, Fusar-Poli P (2021a). Longitudinal outcome of attenuated positive symptoms, negative symptoms, functioning and remission in people at clinical high risk for psychosis: a meta-analysis. eClinicalMedicine 36.

de Pablo GS, Radua J, Pereira J, Bonoldi I, Arienti V, Besana F, Soardo L, Cabras A, Fortea L, Catalan A, Vaquerizo-Serrano J, Coronelli F, Kaur S, Da Silva J, Shin JI, Solmi M, Brondino N, Politi P, McGuire P, Fusar-Poli P (2021b). Probability of Transition to Psychosis in Individuals at Clinical High Risk An Updated Meta-analysis. JAMA Psychiatry 78(9), 970–978.

First MB (2014). Structured Clinical Interview for the DSM (SCID), in: Cautin RL, Lilienfeld SO (Eds.), The Encyclopedia of Clinical Psychology. Wiley Online Library, pp. 1–6.

Fusar-Poli P, Cappucciati M, Borgwardt S, Woods SW, Addington J, Nelson B, Nieman DH, Stahl DR, Rutigliano G, Riecher-Rössler A, Simon AE, Mizuno M, Lee TY, Kwon JS, Lam MML, Perez J, Keri S, Amminger P, Metzler S, Kawohl W, Rössler W, Lee J, Labad J, Ziermans T, An SK, Liu CC, Woodberry KA, Braham A, Corcoran C, McGorry P, Yung AR, McGuire PK (2016a). Heterogeneity of psychosis risk within individuals at clinical high risk: A meta-analytical stratification. JAMA Psychiatry 73(2), 113–120.

Fusar-Poli P, Cappucciati M, Rutigliano G, Lee TY, Beverly Q, Bonoldi I, Lelli J, Kaar SJ, Gago E, Rocchetti M, Patel R, Bhavsar V, Tognin S, Badger S, Calem M, Lim K, Kwon JS, Perez J, McGuire P (2016b). Towards a Standard Psychometric Diagnostic Interview for Subjects at Ultra High Risk of Psychosis: CAARMS versus SIPS. Psychiatry Journal 2016, 7146341.

Fusar-Poli P, Hobson R, Raduelli M, Balottin U (2012). Reliability and validity of the Comprehensive Assessment of the at Risk Mental State, Italian version (CAARMS-I). Current Pharmaceutical Design 18(4), 386–391.

Fusar-Poli P, Salazar de Pablo G, Correll CU, Meyer-Lindenberg A, Millan MJ, Borgwardt S, Galderisi S, Bechdolf A, Pfennig A, Kessing LV, van Amelsvoort T, Nieman DH, Domschke K, Krebs M-O, Koutsouleris N, McGuire P, Do KQ, Arango C (2020). Prevention of Psychosis: Advances in Detection, Prognosis, and Intervention. JAMA Psychiatry 77(7), 755–765.

Grilo CM, Shea MT, Sanislow CA, Skodol AE, Gunderson JG, Stout RL, Pagano ME, Yen S, Morey LC, Zanarini MC, McGlashan TH (2004). Two-year stability and change of schizotypal, borderline, avoidant, and obsessive-compulsive personality disorders. Journal of Consulting and Clinical Psychology 72(5), 767–775.

Hall RC (1995). Global assessment of functioning. A modified scale [see comments]. Psychosomatics 36(3), 267–275.

Hamilton M (1984). Fish’s Schizophrenia. Wright-PSG, Bristol.

Hartmann JA, Schmidt SJ, McGorry PD, Berger M, Berger GE, Chen EYH, de Haan L, Hickie IB, Lavoie S, Markulev C, Mossaheb N, Nieman DH, Nordentoft M, Polari A, Riecher-Rössler A, Schäfer MR, Schlögelhofer M, Smesny S, Thompson A, Verma SK, Yuen HP, Yung AR, Amminger GP, Nelson B (2020). Trajectories of symptom severity and functioning over a three-year period in a psychosis high-risk sample: A secondary analysis of the Neurapro trial. Behaviour Research and Therapy 124, 103527.

Katsura M, Ohmuro N, Obara C, Kikuchi T, Ito F, Miyakoshi T, Matsuoka H, Matsumoto K (2014). A naturalistic longitudinal study of at-risk mental state with a 2.4 year follow-up at a specialized clinic setting in Japan. Schizophrenia Research 158(1-3), 32–38.

Keefe RS, Woods SW, Cannon TD, Ruhrmann S, Mathalon DH, McGuire P, Rosenbrock H, Daniels K, Cotton D, Roy D (2021). A randomized Phase II trial evaluating efficacy, safety, and tolerability of oral BI 409306 in attenuated psychosis syndrome: Design and rationale. Early Intervention in Psychiatry 15(5), 1315–1325.

Kotlicka-Antczak M, Podgorski M, Oliver D, Maric NP, Valmaggia L, Fusar-Poli P (2020). Worldwide implementation of clinical services for the prevention of psychosis: The IEPA early intervention in mental health survey. Early Interv Psychiatry, 1-10.

Kwon JS, Byun MS, Lee TY, An SK (2012). Early intervention in psychosis: Insights from Korea. Asian Journal of Psychiatry 5(1), 98–105.

Lee TY, Lee SS, Gong B-g, Kwon JS (2022). Research Trends in Individuals at High Risk for Psychosis: A Bibliometric Analysis. Frontiers in Psychiatry 13.

Lehembre-Shiah E, Leong W, Brucato G, Abi-Dargham A, Lieberman JA, Horga G, Girgis RR (2017). Distinct Relationships Between Visual and Auditory Perceptual Abnormalities and Conversion to Psychosis in a Clinical High-Risk Population. Jama Psychiatry 74(1), 104–106.

Lho SK, Oh S, Moon SY, Choi W, Kim M, Lee TY, Kwon JS (2021). Reliability and validity of the Korean version of the comprehensive assessment of at-risk mental states. Early Intervention in Psychiatry 15(6), 1730–1737.

Maxwell ME (1992). The Family Interview for Genetic Studies: Manual, National Institute of Mental Health, Washington DC

Mensi MM, Molteni S, Iorio M, Filosi E, Ballante E, Balottin U, Fusar-Poli P, Borgatti R (2021). Prognostic Accuracy of DSM-5 Attenuated Psychosis Syndrome in Adolescents: Prospective Real-World 5-Year Cohort Study. Schizophrenia Bulletin 47(6), 1663–1673.

Miller TJ, McGlashan TH, Woods SW, Stein K, Driesen N, Corcoran CM, Hoffman R, Davidson L (1999). Symptom assessment in schizophrenic prodromal states. Psychiatr Q 70(4), 273–287.

Miyakoshi T, Matsumoto K, Ito F, Ohmuro N, Matsuoka H (2009). Application of the comprehensive assessment of atLJrisk mental states (CAARMS) to the Japanese population: Reliability and validity of the Japanese version of the CAARMS. Early Intervention in Psychiatry 3(2), 123–130.

Morosini PL, Magliano L, Brambilla La, Ugolini S, Pioli R (2000). Development, reliability and acceptability of a new version of the DSMLJIV Social and Occupational Functioning Assessment Scale (SOFAS) to assess routine social funtioning. Acta Psychiatrica Scandinavica 101(4), 323–329.

Morrison AP, French P, Stewart SLK, Birchwood M, Fowler D, Gumley AI, Jones PB, Bentall RP, Lewis SW, Murray GK, Patterson P, Brunet K, Conroy J, Parker S, Reilly T, Byrne R, Davies LM, Dunn G (2012). Early detection and intervention evaluation for people at risk of psychosis: Multisite randomised controlled trial. BMJ (Online) 344(7852).

Mourgues C, Benrimoh D, Addington J, Bearden C, Cadenhead K, Tsuang M, Cornblatt B, Keshavan M, Stone W, Mathalon D, Perkins D, Walker E, Cannon T, Woods S, Shah J, Powers A (2023). Emergence of Delusions and Hallucinations in High-Risk and First-Episode Samples, Schizophrenia International Research Society, Toronto,

Nelson B, Yuen HP, Amminger GP, Berger G, Chen EYH, de Haan L, Hartmann JA, Hickie IB, Lavoie S, Markulev C, Mossaheb N, Nieman DH, Nordentoft M, Polari A, Riecher-Rössler A, Schäfer MR, Schlögelhofer M, Smesny S, Tedja A, Thompson A, Verma S, Yung AR, McGorry PD (2022). Distress Related to Attenuated Psychotic Symptoms: Static and Dynamic Association With Transition to Psychosis, Nonremission, and Transdiagnostic Symptomatology in Clinical High-Risk Patients in an International Intervention Trial. Schizophrenia Bulletin Open 3(1), sgaa006.

Niles HF, Walsh BC, Woods SW, Powers III AR (2019). Does hallucination perceptual modality impact psychosis risk? Acta Psychiatrica Scandinavica 140(4), 360–370.

Oliver D, Arribas M, Radua J, Salazar de Pablo G, De Micheli A, Spada G, Mensi MM, Kotlicka-Antczak M, Borgatti R, Solmi M, Shin JI, Woods SW, Addington J, McGuire P, Fusar-Poli P (2022). Prognostic accuracy and clinical utility of psychometric instruments for individuals at clinical high-risk of psychosis: a systematic review and meta-analysis. Molecular Psychiatry 27(9), 3670–3678.

Olsen KA, Rosenbaum B (2006). Prospective investigations of the prodromal state of schizophrenia: Assessment instruments. Acta Psychiatrica Scandinavica 113(4), 273–282.

Paterlini F, Pelizza L, Galli G, Azzali S, Scazza I, Garlassi S, Chiri LR, Poletti M, Pupo S, Raballo A (2019). Interrater reliability of the authorized Italian version of the Comprehensive Assessment of At-Risk Mental States (CAARMS-ITA). Journal of Psychopathology 25, 24–28.

Perkins DO, Jeffries CD, Cornblatt BA, Woods SW, Addington J, Bearden CE, Cadenhead KS, Cannon TD, Heinssen R, Mathalon DH, Seidman LJ, Tsuang MT, Walker EF, McGlashan TH (2015). Severity of thought disorder predicts psychosis in persons at clinical high-risk. Schizophrenia Research 169(1-3), 169–177.

Power L, Polari AR, Yung AR, McGorry PD, Nelson B (2016). Distress in relation to attenuated psychotic symptoms in the ultra-high-risk population is not associated with increased risk of psychotic disorder. Early Intervention in Psychiatry 10(3), 258–262.

Pratt DN, Bridgwater M, Schiffman J, Ellman LM, Mittal VA (in press). Do the Components of Attenuated Positive Symptoms Truly Represent One Construct? Schizophrenia Bulletin, sbac182 https://doi.org/110.1093/schbul/sbac1182.

Rapado-Castro M, McGorry PD, Yung A, Calvo A, Nelson B (2015). Sources of clinical distress in young people at ultra high risk of psychosis. Schizophr Res 165(1), 15–21.

Rekhi G, Rapisarda A, Lee J (2019). Impact of distress related to attenuated psychotic symptoms in individuals at ultra high risk of psychosis: Findings from the Longitudinal Youth at Risk Study. Early Intervention in Psychiatry 13(1), 73–78.

Salazar de Pablo G, Catalan A, Fusar-Poli P (2020). Clinical Validity of DSM-5 Attenuated Psychosis Syndrome: Advances in Diagnosis, Prognosis, and Treatment. JAMA Psychiatry 77(3), 311–320.

Salazar de Pablo G, Woods SW, Drymonitou G, de Diego H, Fusar-Poli P (2021). Prevalence of Individuals at Clinical High-Risk of Psychosis in the General Population and Clinical Samples: Systematic Review and Meta-Analysis. Brain Sciences 11(11), 1544.

Schultze-Lutter F, Michel C, Ruhrmann S, Schimmelmann BG (2014). Prevalence and clinical significance of DSM-5-attenuated psychosis syndrome in adolescents and young adults in the general population: The Bern Epidemiological At-Risk (BEAR) Study. Schizophrenia Bulletin 40(6), 1499–1508.

Schultze-Lutter F, Schimmelmann BG, Ruhrmann S (2011). The Near Babylonian Speech Confusion in Early Detection of Psychosis. Schizophrenia Bulletin 37(4), 653–655.

Schultze-Lutter F, Schimmelmann BG, Ruhrmann S, Michel C (2013). ’A rose is a rose is a rose’, but at-risk criteria differ. Psychopathology 46(2), 75–87.

Segal JH (1989). Erotomania revisited - From Kraepelin to DSM-III-R. American Journal of Psychiatry 146(10), 1261–1266.

Sheehan DV, Lecrubier Y, Sheehan KH, Amorim P, Janavs J, Weiller E, Hergueta T, Baker R, Dunbar GC (1998). The Mini-International Neuropsychiatric Interview (M.I.N.I.): The development and validation of a structured diagnostic psychiatric interview for DSM-IV and ICD-10. Journal of Clinical Psychiatry 59(SUPPL. 20), 22-33.

Smeets F, Lataster T, Viechtbauer W, Delespaul P, Group (2015). Evidence That Environmental and Genetic Risks for Psychotic Disorder May Operate by Impacting on Connections Between Core Symptoms of Perceptual Alteration and Delusional Ideation. Schizophenia Bulletin 41(3), 687–697.

U.S. Department of Health and Human Services, Food and Drug Administration, Center for Drug Evaluation and Research (CDER), Center for Biologics Evaluation and Research (CBER), Center for Devices and Radiological Health (CDRH) (2022). Patient-Focused Drug Development: Selecting, Developing, or Modifying Fit-for-Purpose Clinical Outcome Assessments Guidance for Industry, Food and Drug Administration Staff, and Other Stakeholders, https://www.fda.gov/regulatory-information/search-fda-guidance-documents/patient-focused-drug-development-selecting-developing-or-modifying-fit-purpose-clinical-outcome.

Walsh BC (2021). SIPS Certified Assessors and Training. https://thesipstraining.com.

Wang P, Yan CD, Dong XJ, Geng L, Xu C, Nie Y, Zhang S (2022). Identification and predictive analysis for participants at ultra-high risk of psychosis: A comparison of three psychometric diagnostic interviews. World Journal of Clinical Cases 10(8), 2420–2428.

Wilson RS, Shryane N, Yung AR, Morrison AP (2020). Distress related to psychotic symptoms in individuals at high risk of psychosis. Schizophrenia Research 215, 66–73.

Woods SW, Choi J, Mamah D (2021). Full speed ahead on indicated prevention of psychosis. World Psychiatry 20(2), 223.

Woods SW, Miller TJ, McGlashan TH (2001). The “prodromal” patient: both symptomatic and at-risk. CNS Spectr 6(3), 223–232.

Woods SW, Walsh BC, Addington J, Cadenhead KS, Cannon TD, Cornblatt BA, Heinssen R, Perkins DO, Seidman LJ, Tarbox SI, Tsuang MT, Walker EF, McGlashan TH (2014). Current status specifiers for patients at clinical high risk for psychosis. Schizophrenia Research 158, 69–75.

Woods SW, Walsh BC, Powers III AR, McGlashan TH (2019). Reliability, validity, epidemiology, and cultural variation of the Structured Interview for Psychosis-risk Syndromes (SIPS) and the Scale Of Psychosis-risk Symptoms (SOPS), in: Li H, Shapiro DI, Seidman LJ (Eds.), Handbook of Attenuated Psychosis Syndrome Across Cultures: International Perspectives on Early Identification and Intervention. Springer, New York, pp. 85–113.

Yokusoglu C, Ercis M, Caglar N, Aydemir O, Ucok A (2021). Reliability and validity of the Turkish version of comprehensive assessment of at risk mental states. Early Intervention in Psychiatry 15(4), 1028–1032.

Yung A, Parker S, Davies K, Lin A, Shone C (2015). Comprehensive Assessment of At Risk Mental States, University of Manchester.

Yung AR, McGorry PD, McFarlane CA, Jackson HJ, Patton GC, Rakkar A (1996). Monitoring and care of young people at incipient risk of psychosis. Schizophrenia Bulletin 22(2), 283–303.

Yung AR, Yuen HP, McGorry PD, Phillips LJ, Kelly D, Dell’Olio M, Francey SM, Cosgrave EM, Killackey E, Stanford C, Godfrey K, Buckby J (2005). Mapping the onset of psychosis: The comprehensive assessment of at-risk mental states. Australian & New Zealand Journal of Psychiatry 39(11-12), 964–971.

Zhang TH, Xu LH, Tang YY, Cui HR, Wei YY, Tang XC, Hu Q, Wang Y, Zhu YK, Jiang LJ, Hui L, Liu XH, Li CB, Wang JJ (2018). Isolated hallucination is less predictive than thought disorder in psychosis: Insight from a longitudinal study in a clinical population at high risk for psychosis. Scientific Reports 8(1).

