## Appendix S1 for "Development of the PSYCHS: Positive SYmptoms and Diagnostic Criteria for the CAARMS Harmonized with the SIPS"

**Appendix S1.** Supporting information

**PSYCHS Psychosis and Clinical High Risk (CHR)/Ultra-High Risk (UHR) Syndrome Categories**

**Harmonized Criteria for CAARMS/SIPS Psychosis Diagnosis**

- A psychotic severity/intensity symptom rating=6 on at least one of P1-P15

**AND EITHER**

- Symptom lasts >=1 week at severity/intensity=6 and frequency >=4 (3-6 days/wk – more than 1 hr/day **OR** daily < 1 hr/day) **UNLESS** truncated by new or increased antipsychotic treatment

**OR**

- Symptom while rated=6 was imminently dangerous (physically or to personal dignity or to social/family networks).

| **Modified SIPS CHR Lifetime Syndrome Criteria** | | |
| --- | --- | --- |
| **Brief Intermittent Psychotic Syndrome (BIPS)** | **Attenuated Positive Symptom Syndrome (APSS)** | **Genetic Risk and Deterioration (GRD)** |
| - A psychotic severity/intensity symptom (rating= 6) on at least one of P1-P15 - Present at least **several minutes a day**, but has not lasted >=1 week at severity/intensity=6 and frequency >=4 (3-6 days/wk – more than 1 hr/day; OR daily < 1 hr/day) - Symptoms rated a 6 are not imminently dangerous (physically or to personal dignity or to social/family networks) - Not better explained by another DSM disorder | - At least one of P1-P15 rated severity/intensity **3, 4 or 5** - Symptom must occur at an average frequency of **at least once per week over a month** - Not better explained by another DSM-5 disorder   **Indicate whether symptoms were sufficiently distressing and disabling to the participant to warrant clinical attention | Family history of psychosis in **first degree relative** OR S**chizotypal Personality Disorder** in identified participant  **AND**  **Drop in functioning:**   - Impact: SOFAS score over any month **at least 30% below** previous level of functioning over the month one year earlier |

| **SIPS BIPS Current Status**  (requires that Lifetime criteria have been met) | | | |
| --- | --- | --- | --- |
| **Progression** | **Persistence** | **Partial Remission** | **Full Remission** |
| BIPS qualifying symptoms occur at severity/intensity=6 at least several minutes per day at least one day in the past month **AND** began or worsened to a severity/intensity =6 in the past three months | BIPS qualifying symptoms occur at severity/intensity=6 but **did not** begin in the past 3 months | - **First Pathway**: BIPS qualifying symptom  previously rated severity/intensity=6 now currently rated severity/intensity <=5 for six months or less (i.e., met severity/intensity rating 6 within the past 6 months, but not recently within the past month) - **Second Pathway**: previously qualifying lifetime symptoms rated severity/intensity=6 now do not occur at least several minutes per day at least once in the past month or are now better explained by another DSM disorder | Previously BIPS qualifying symptoms currently score severity/intensity <=5 and for more than six months. |

| **SIPS APSS Current Status**  (requires that Lifetime criteria have been met) | | | |
| --- | --- | --- | --- |
| **Progression** | **Persistence** | **Partial Remission** | **Full Remission** |
| Qualifying symptoms began **within the past year** OR currently rate one or more scale points higher **compared to 12 months ago** | Qualifying symptoms **did not** begin within the past year **AND do not** currently rate one or more scale points higher compared to 12 months ago | - **First Pathway**: previously qualifying symptom now currently rated severity/intensity <=2 for six months or less (i.e., met severity/intensity rating 3-5 within the past 6 months but not recently within the past month) - **Second Pathway**: previously qualifying symptoms now do not occur at an average frequency of at least once per week over the past month or are now better explained by another DSM disorder | Previously qualifying symptoms currently score **<=2** for more than six months |

| **Modified SIPS GRD Current Status**  (requires that Lifetime criteria have been met) | | | |
| --- | --- | --- | --- |
| **Progression** | **Persistence** | **Partial Remission** | **Full Remission** |
| SOFAS score over past month **at least 30% below** previous level of functioning over the month one year earlier | SOFAS score over past month **is less than 30% below** previous level of functioning over the month one year earlier but also lower than 90% of the premorbid level | - SOFAS score over past month **is at least**  90% of the premorbid level but for six months or less (i.e., SOFAS was <90% of premorbid within the past 6 months but not recently over the past month) | SOFAS score **currently at least** 90% of the premorbid level for more than six months |

| **Modified CAARMS UHR Syndromes** | | |
| --- | --- | --- |
| **Brief Limited Intermittent Psychotic Symptoms (BLIPS)** | **Attenuated Positive Symptoms (APS) – Subthreshold Intensity OR Frequency** | **Vulnerability** |
| - Symptoms present **OVER THE PAST YEAR** of psychotic severity/intensity **= 6** on any P1-P15   **AND**   - Symptom frequency rating is **>=4** (i.e., 3-6 days/wk - >1 hour/day; or daily - <1 hour/day)   **AND**   - **Duration < 7 days** (with spontaneous remission each time)   **Note: while at severity/intensity = 6, symptoms CAN NOT occur ONLY during peak intoxication from hallucinogens, amphetamines or cocaine (disregard alcohol and cannabis). | Symptoms present for **OVER THE PAST YEAR** at EITHER:   - **Subthreshold Intensity:** Severity/Intensity rating of **3-5** on any P1-P15 **AND** symptom frequency rating is **>=3** (i.e., 1 day/month to 2 days/wk – >1 hour/day; OR 3-6 days/wk - <1 hour/day)   **OR**   - **Subthreshold Frequency:** Severity/Intensity rating of **6** on any P1-P15 **AND** symptom frequency rating = **3** (i.e., 1 day/month to 2 days/wk – >1 hour a day; OR 3-6 days/wk - <1 hour/day.)   **Note: while at highest severity/intensity, symptoms CAN NOT occur ONLY during peak intoxication from hallucinogens, amphetamines or cocaine (disregard alcohol and cannabis). | Family history of psychosis in **first degree relative** OR S**chizotypal Personality Disorder** in identified participant  **AND EITHER**  **Drop in functioning:**   - Impact: SOFAS score **at least 30% below** previous level of functioning and sustained for **at least one month** - Recency: Change in functioning occurred **within last year**   **OR**  **Sustained low functioning:**   - Impact: SOFAS score of 50 or less] - Recency: For the **past 12 months** or longer |
